# Spatial modeling cannot currently differentiate SARS-CoV-2 coronavirus and human distributions on the basis of climate in the United States

**DOI:** 10.1101/2020.04.08.20057281

**Authors:** Robert Harbert, Seth W. Cunningham, Michael Tessler

**Affiliations:** Stonehill College, Easton, Massachusetts; Sackler Institute for Comparative Genomics, American Museum of Natural History, New York, New York

**Keywords:** climate, coronavirus, COVID-19, SARS-CoV-2, species distribution modeling, US

## Abstract

The SARS-CoV-2 coronavirus is wreaking havoc globally, yet knowledge of its biology is limited. Climate and seasonality influence the distributions of many diseases, and studies suggest a link between SARS-CoV-2 and cool weather. One such study, building species distribution models (SDMs), predicted SARS-CoV-2 risk may remain concentrated in the Northern Hemisphere, shifting northward in summer months. Others have highlighted issues with SARS-CoV-2 SDMs, notably: the primary niche of the virus is the host it infects, climate may be a weak distributional predictor, global prevalence data have issues, and the virus is not in a population equilibrium. While these issues should be considered, climate still may be important for predicting the future distribution of SARS-CoV-2. To further examine if there is a link, we model with raw cases and population scaled cases for SARS-CoV-2 county-level data from the United States. We show that SDMs built from population scaled cases data cannot be distinguished from control models built from raw human population data, while SDMs built on raw data fail to predict the current known distribution of cases in the US. The population scaled analyses indicate that climate may not play a central role in current US viral distribution and that human population density is likely a primary driver. Still, we do find slightly more population scaled viral cases in cooler areas. This coupled with our geographically constrained focus make it so we cannot rule out climate as a partial driver of the US SARS-CoV-2 distribution. Climate’s role on SARS-CoV-2 should continue to be cautiously examined, but at this time we should assume that SARS-CoV-2 can spread anywhere in the US.

## Introduction

### SARS-CoV-2 and climate overview

As of March 26, 2020, the United States of America has had the most reported cases of COVID-19, the disease caused by the coronavirus SARS-CoV-2 (CDC NCIRD, 2020). While we are beginning to better understand the global and US-level distributions of this virus, we are only starting to learn how abiotic variables affect its basic distribution. With over two months of SARS-CoV-2 records across US counties, the role of abiotic variables is becoming easier to examine, especially as multiple sources are compiling data and making it public (Dong et al., 2020; The COVID Tracking Project, 2020; The New York Times, 2020).

Organisms are distributed within their environment based on both direct and indirect interactions with biotic and abiotic variables (Elith & Leathwick, 2009) — SARS-CoV-2 is no exception. As this virus is primarily distributed by human hosts, it is constrained by human distributions and interactions (i.e., biotic variables). There are numerous documented cases on local and global scales of individual human sources for new outbreaks (Holshue et al., 2020; KCDC, 2020). Still, the SARS-CoV-2 has thus far been most concentrated in the Northern Hemisphere; even there, to date, it has only proliferated in a subset of population centers. Furthermore, many other viruses, including coronaviruses, display marked seasonality and are affected by local climatic conditions (Fisman, 2012; Gaunt et al., 2010; Lofgren et al., 2007; Price et al., 2019). This has prompted researchers to begin looking globally at how weather and climate (i.e., a subset of abiotic variables) may relate to the presence and abundance of the virus.

Modeling the SARS-CoV-2 viral distribution in relation to climate and other abiotic variables could help refine our understanding, hopefully adding to the knowledge gleaned from studies on human transmission dynamics (Chinazzi et al., 2020; Kucharski et al., 2020), which appear to be explanatory for much of the viral distribution. Climate can also be compared directly to biotic variables, such as human population density, to disentangle their relative importances. More dense human populations can have elevated communicable disease spread, as there is more person-to-person contact (Fang et al., 2013) — the principal way SARS-CoV-2 is known to spread (Rothan & Byrareddy, 2020). Any improved understanding of the abiotic variables influencing the niche of SARS-CoV-2 might, in part, help prevent viral spread. However, it is important to make clear that the models presented here, and in other recent studies (Araújo & Naimi, 2020; Bariotakis et al., 2020), are influenced by the variability of input parameters, the variables explored, and the restrictions inherent in modeling of any complex system (Elith et al., 2011). We present this study to serve as a baseline in our understanding, given the available data, rather than a definitive model, and hope that our work acts as a jumping off point for other studies.

### SARS-CoV-2 and climate research

Early studies suggest that transmission of SARS-CoV-2 may have, at minimum, a loose association with climatic features. There have been numerous reports showing a correlation of case incidence with cool temperatures and low humidity (Alvarez-Ramirez & Meraz, 2020; Araújo & Naimi, 2020; Bannister-Tyrrell et al., 2020; Chen et al., 2020; Ficetola & Rubolini, 2020; Notari, 2020; Wang et al., 2020). Furthermore, studies that controlled for case growth rate or demographic factors found that weather was still a significant factor in the success of SARS-CoV-2 outbreaks (Bukhari & Jameel, 2020; Chen et al., 2020; Sajadi et al., 2020). Unfortunately, none of these studies provide a mechanism for how abiotic variables like temperature might limit or promote the person-to-person transmission of the virus. Still, lipid enveloped viruses (including coronaviruses) may be more stable outside of the host in lower humidity and cooler temperatures, conditions that are common in temperate areas in late winter and early spring (Price et al., 2019). Furthermore, a mechanism could be indirect, as humans behave differently in different seasons (Bedford et al., 2015; Wesolowski et al., 2017).

Scientists often employ species distribution modeling (SDM; sometimes called ecological niche modeling) to predict geographic ranges of species. SDMs employ environmental data (typically climate) to predict if geographic space is suitable for a given species or population (Peterson, 2001). These models have proven useful in a wide variety of applications, such as invasion biology, climate change, zoonotic diseases, and speciation (Elith & Leathwick, 2009; Guisan & Thuiller, 2005).

Researchers have begun to create SDMs of SARS-CoV-2 using climatic variables (Araújo & Naimi, 2020; Bariotakis et al., 2020). These studies suggest that the virus is strongly constrained by global climate patterns (Araújo & Naimi, 2020; Bariotakis et al., 2020). Preliminary models also suggest that the virus will continue to be concentrated in the Northern Hemisphere, shifting northwards throughout the summer and then back towards its current distribution in the fall and winter (Araújo & Naimi, 2020).

However, a research group recently put forth a strong rebuttal (Chipperfield et al., 2020) to some of this SDM work (Araújo & Naimi, 2020). Their main criticism asserts that basic assumptions underlying SDMs are violated when modeling SARS-CoV-2, due to the mode of transmission, current population disequilibrium, and failure to incorporate epidemiological data (Chipperfield et al., 2020). Furthermore, they highlight issues pertaining to input data for global records, such as: Missing data, omitted data, and single localities representing many thousands of reported cases (hospital coordinates or political centroids) across an entire country (Araújo et al., 2019). This is exacerbated as many countries are reporting records based on varied criteria, such as: Multiple molecular tests and tests using computed tomography scans (Lippi et al., 2020). The rebuttal additionally suggested that model evaluation and justification were insufficient. An updated draft of the original paper (Araújo & Naimi, 2020) has addressed some of these issues, and produced reasonable arguments to some others. While we agree with many arguments from the rebuttal, we believe SDMs have the potential to be useful for modeling in a number of instances, if done carefully, and have been effective when used for other diseases (Carlson et al., 2016; Pigott et al., 2014, 2015) and other instances when data are limited or incomplete (Barbet-Massin et al., 2018; Fois et al., 2018; Galante et al., 2018; Hernandez et al., 2006; Katz & Zellmer, 2018; Kiedrzyński et al., 2017; Pearson et al., 2006).

Still, the current climate SDMs of SARS-CoV-2 may simply reflect the habitat preferences of their host, humans. Careful consideration of host availability (human population density) and pathogen ecologies (abiotic variables related to transmission) may be necessary to frame analyses modeling the global distribution (Johnson et al., 2019), helping to better ensure that projected distributions are not simply the result of environmental variables related to human population density.

### SARS-CoV-2 in the US

COVID-19 was first detected in the US on January 20, 2020 (Holshue et al., 2020), has since been detected in all 50 states, and continues to spread rapidly (Chinazzi et al., 2020). Quality county-level data are also publically available (Dong et al., 2020; The COVID Tracking Project, 2020; The New York Times, 2020). SARS-CoV-2 is predicted, as of April 4, 2020, to cause >90,000 deaths in the US (IHME COVID-19 team & Murray, 2020). However, like much of the world, the case distribution is rapidly changing and the case numbers are rapidly growing. Models and projections are clearly subject to change as incoming case data increases. Public policy decisions (e.g., self isolation) are also surely going to impact the spread of this coronavirus, and are difficult to account for at the time due to the lag time between policy and changes in case load.

While our models and the models produced elsewhere are sure to have issues, COVID-19 may peak in the US largely throughout April (IHME COVID-19 team & Murray, 2020). Accordingly, the time to produce predictions is now. We are creating a baseline of information that can, and should be, evaluated and tested as more data become available in more regions. Ultimately, these baselines may be particularly helpful not only in the US, but in other countries that are only just beginning to encounter the pandemic.

## Methods

### Code and results deposition

The code and results used in this study are deposited under a CC-BY-NC 4.0 License on Github (https://github.com/rsh249/cv19_enm/releases/tag/v0.0.2). All analysis code was written using R 3.6.2 (R Core Team, 2019). Plots for Figures 1, 2, 3, S1, and S2 produced using the “ggplot2” package (Wickham, 2009).

**Figure 1.**
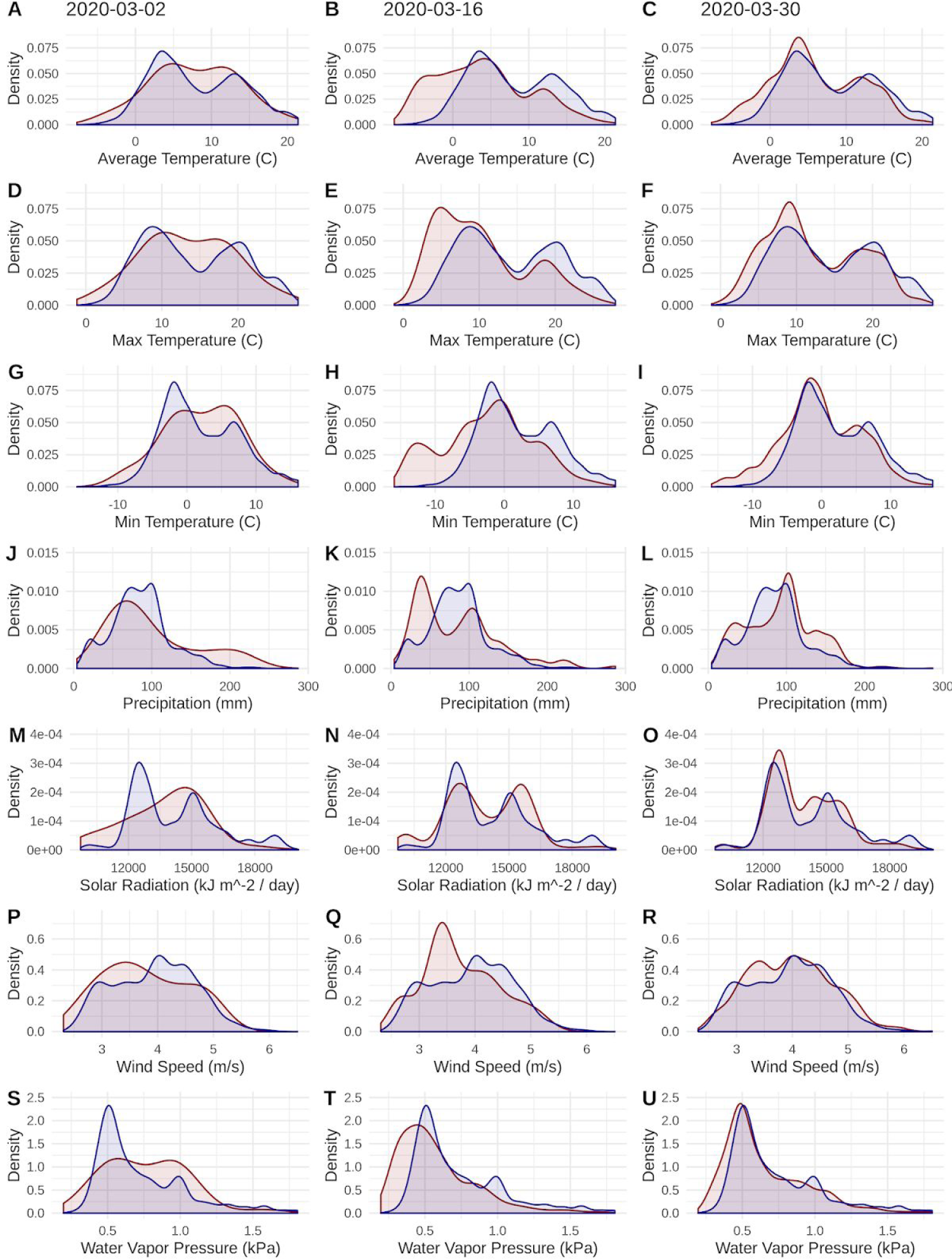
Probability densities of SARS-CoV-2 coronavirus cases (using population scaled data; curves in red) compared to the probability densities of human populations (curves in blue) in each US county for each of seven climate variables. Probability density curves are standardized to an area of one.

**Figure 2.**
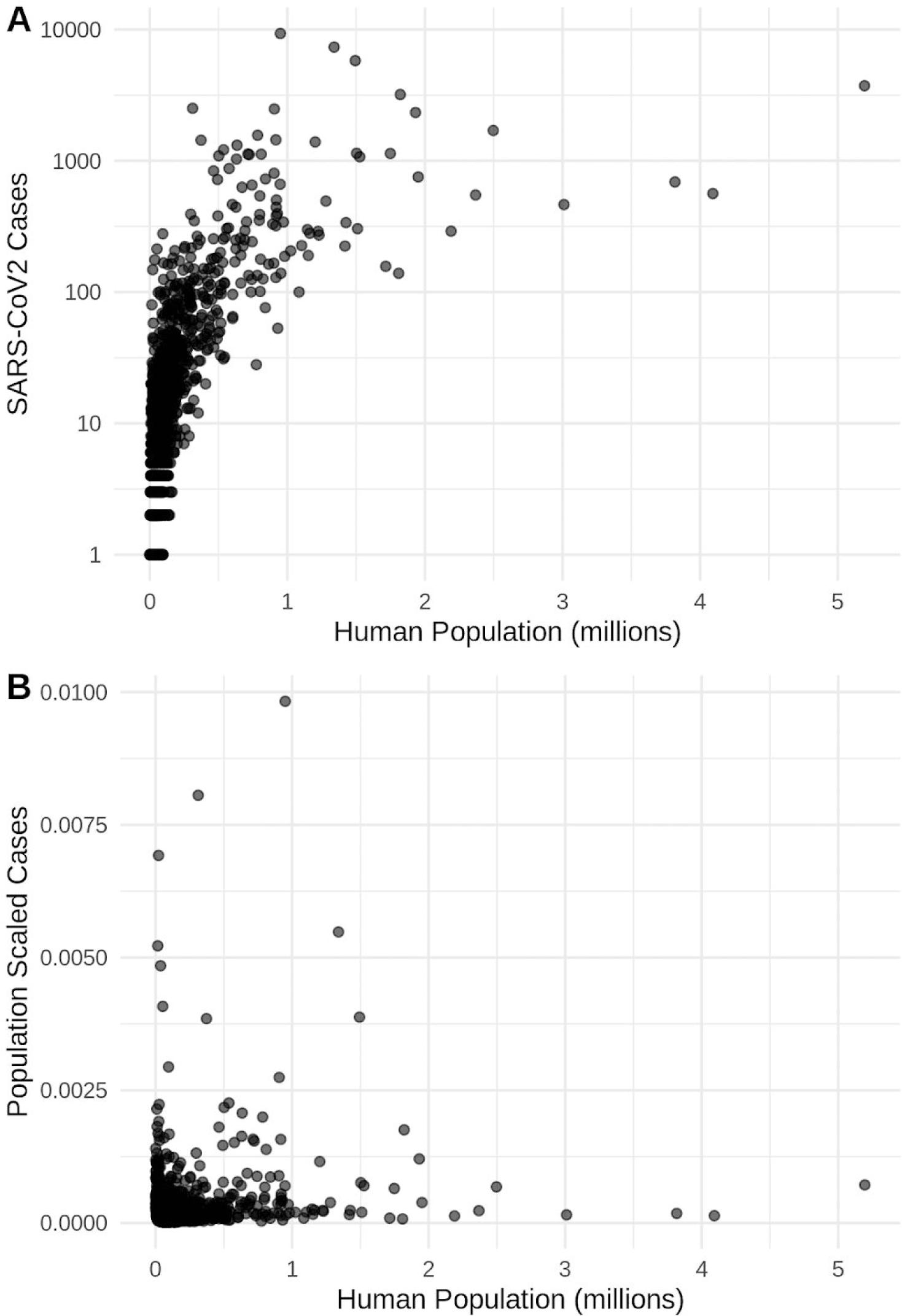
The relationship in the US between human population size and SARS-CoV-2 coronavirus cases, using (A) total viral cases and (B) population scaled viral cases. New York City, an outlying point, has been excluded for clearer visualization.

**Figure 3.**
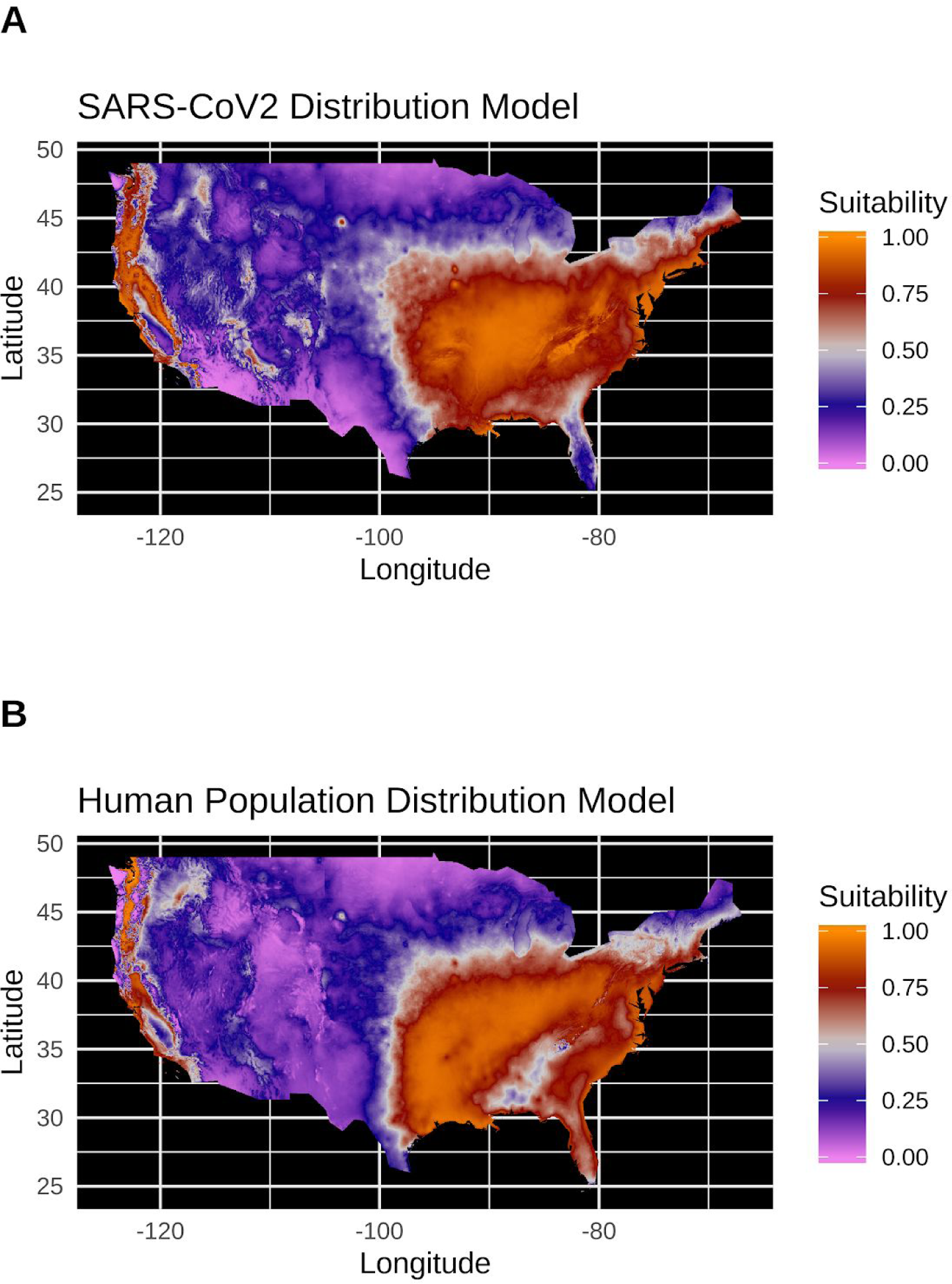
(A) Species distribution model of the SARS-CoV-2 coronavirus (using population scaled data) for March 30, 2020. (B) Human population distribution model for the US from 2010.

### Data acquisition

SARS-CoV-2 case data for US counties were collected from the New York Times database (The New York Times, 2020) on March 31, 2020 for the March 30, 2020 data release. County-level data on human population densities were acquired from the 2010 United States Census through the R “tidycensus” package (Walker et al., 2020). Georeferencing to county centers was performed by referencing county and state names in the GeoNames database (https://www.geonames.org/). While exact virus case geolocations or even town-level data would provide finer scale resolution to our analyses, these data are likely the best curated dataset for the US at this time.

### Climate data

Climate data averaged from 1970-2000 for the month of March were accessed through the WorldClim v2.1 database (Fick & Hijmans, 2017). The seven climate parameters examined were average temperature, minimum temperature, maximum temperature, average precipitation, solar radiation, wind speed, and water vapor pressure (a measure related to humidity). Climate values were extracted from these layers for each georeferenced county record using the average from a 5km buffer around each county center coordinate. While county centers may not be ideal in all circumstances, our view is that county centers plus this generous buffer will average the climate relative to most cases in the area. WIthout high levels of individual case and movement tracking, more precise georeferencing of viral transmission is not currently possible.

### Basic examinations

Data for SARS-CoV-2 cases and human population totals were each compared to the seven abiotic variables listed above. Probability densities are calculated from occurrence and corresponding climate data using a Gaussian Kernel Density Estimator and standard bandwidth. Raw case data, total cases per county, were scaled to reflect virus cases in each county unit by dividing total cases by the county population from the 2010 US Census. These population scaled viral cases (cases / population) were used as probability density weightings, and resulting curves were standardized to an area of one. Probability densities were also calculated with raw case count data and county-level population as weightings. Probability density estimation and visualization was done with the R ‘ggplot2’ package (Wickham, 2009). To test whether the SARS-CoV-2 cases and human population data were correlated, we applied Spearman tests “cor.test(method=‘spearman’)”.

### Species distribution models

Occurrence data for raw SDMs were generated by expanding each climate record by the total SARS-CoV-2 case count for each county. Occurrence data for population scaled SDMs were generated by expanding each record used in the probability density estimation such that no county had fewer than one record [i.e., cases / population × 100,000 (the expansion factor for our data)]. Maxent model parameters were tested by building a suite of SDMs using the ENMeval package (Muscarella et al., 2014) for SARS-CoV-2 distribution data with occurrences generated from raw reported virus case values and population scaled case values. Operational models using the best model parameters (identified by minimizing AICc) were then built for SARS-CoV-2 using all population scaled data, raw virus data, and for human density using the county population data with the Maxent algorithm implemented in the “maxnet” R package (Phillips, 2017). Of the seven WorldClim variables, the three temperature variables are clearly linked. Accordingly, we only used maximum temperature, as higher temperatures have been hypothesized to lower the viral distribution. Keeping all temperature variables made similar models, but are not further reported on, as reducing correlated variables is generally preferable (Araújo et al., 2019).

Given recent critiques, it has become clear that when using SDMs in relation to SARS-CoV-2 it is worth documenting how close a study may come to the, in our opinion somewhat asparational, gold standards for this type of modeling (Araújo et al., 2019). The mode rank for our SDMs is bronze (**Table 1**), largely as data from an emerging disease are inherently imperfect.

**Table 1.** Evaluation of our SDM practices against the best practices that have been proposed for this field (Araújo et al., 2019).

| Guideline | Standard | Justification |
| --- | --- | --- |
| Response variables | A) Sampling: Bronze | Best data available; municipalities, local governments, and states choose who to test. Positive tests only reported. |
|  | B) Identification: Gold | Assuming best practices in testing and reporting. |
|  | C) Spatial accuracy: Bronze | County assignments provide a rough georeference for each record, but do not precisely describe where transmission of the virus occurred. Spatial accuracy unknown. Occurrences limited to identifiable county level localities. |
|  | D) Environmental extent: Deficient | Limiting the study area to the continental US is unlikely to adequately test environmental boundaries. |
|  | E) Geographic extent: Bronze | Study area to include current range in the US. |
| Predictor variables | A) Selection of candidates: Bronze/Deficient | Unclear and not well documented correlations between SARS-CoV-2 transmission and climate. At best, distal variables with weak, indirect control on the distribution. |
|  | B) Spatial and temporal resolution: Deficient | Variables sampled from a 2.5 arcminute grid for all cells within 5km of each occurrence point. Mean value used for modeling. Monthly climate averages as predictors for end of March occurrence data. |
|  | C) Uncertainty: Bronze | Temporal and spatial uncertainty in occurrence data has unquantified potential effects on the model output. |
| Model building | A) Model Complexity: Silver | ENMeval for model testing and selection (AICc) using internal cross validation through the block resampling method. |
|  | B) Treatment of response bias: Silver | Internal cross validation to evaluate bias effects in different models. |
|  | C) Treatment of collinearity: Bronze | “Approximate methods are applied” — Predictor variables hand selected from monthly climate data available to avoid collinearity (i.e., used only Tmax and not Tavg or Tmin). |
|  | D) Uncertainty: Bronze | Multiple Maxent model parameters tested, but only the best model presented. |
| Model evaluation | A) Evaluation of model assumptions: Gold/Silver | Select robust models from all tested models with ENMeval. |
|  | B) Evaluation of model outputs: Silver | Evaluated against multiple, non-independent, geographically structured sub-samples. |
|  | C) Measures of model performance: Silver | Suite of model performance metrics performed via ENMeval. |
| **SUMMARY** | **Mode of the scores: Bronze** | **Model building and testing is generally robust, but data and geographic scope are incomplete at this time.** |

Niche overlap and similarity tests were conducted with the “ecospat” library (Broennimann et al., 2012; Di Cola et al., 2017) to compare the climatic niche of SARS-CoV-2 and humans, as a significant overlap between these would indicate that the virus distribution is not necessarily constrained by climate (i.e., that the full range of human occupied climate is accessible to the virus).

## Results

### Basic examinations

The human population climate curves are visually similar compared to the population scaled SARS-CoV-2 climate curves for all seven abiotic variables during the most recent record in March (**Figure 1**). There do appear to be modest differences in the population scaled viral curves towards cooler temperatures and lower water vapor pressure compared to the human population curves. The raw density of SARS-CoV-2 cases (**Figure S1**) is visually different from the population scaled data (**Figure 1**). Population scaled SARS-CoV-2 data appear to become better fitted to the human population data over time (i.e., these curves better match on March 30 than they did on March 2 or 16, 2020; **Figure 1**), whereas the raw SARS-CoV-2 case data become more and more fixed on a narrow range of values for all climate variables (**Figure S1)**. The latter (fixed and narrow) range presumably has to do with a few virus hotspots.

There is a highly significant positive relationship (Spearman’s *r* = 0.75, *p* < 2e^−12^); between the number of humans in a county and the number of viral cases; however, the population scaled viral cases do not have a strong relationship (Spearman’s *r* = 0.05, *p* = 0.03) with human populations (**Figure 2**). Although the latter *p*-value is significant, this is often not all that meaningful for large datasets, whereas the weak *r* value better illustrates the relationship for the population scaled data.

### Species distribution models

SDMs for SARS-CoV-2 (population scaled cases) and human populations appear to be highly similar, with areas of high suitability predicted for much of the west coast and most of the eastern half of the US (**Figure 3**). There are some differences; for instance, the map for the virus suggests Florida is less suitable than it is in the map for people. The SDM of the raw virus cases (**Figure S2**) fails to reconstruct the known distribution of viral cases in much of the US, and produces a divergent model from the population scaled model. The SDM for humans (**Figure 3, S2**) does not exactly match current human distribution for the US. This was expected, because the goal of the model is to match climates that are correlated with the places where most people live (i.e., it is not simply a map of current human populations). We did this as our ultimate goal was comparing a viral model to a human model.

The niche overlap and similarity tests both find that the models of SARS-CoV-2 (population scaled viral case data) and human population have significantly higher overlap (*p* < 0.01) than would be expected by chance (**Figure 4**), confirming the visual similarities between these maps. The raw data showed less overlap: The similarity test was significant, but an order of magnitude less so than for the population scaled data (*p* = 0.02), while the overlap test was insignificant (*p* = 0.20), as seen in **Figure S3**.

**Figure 4.**
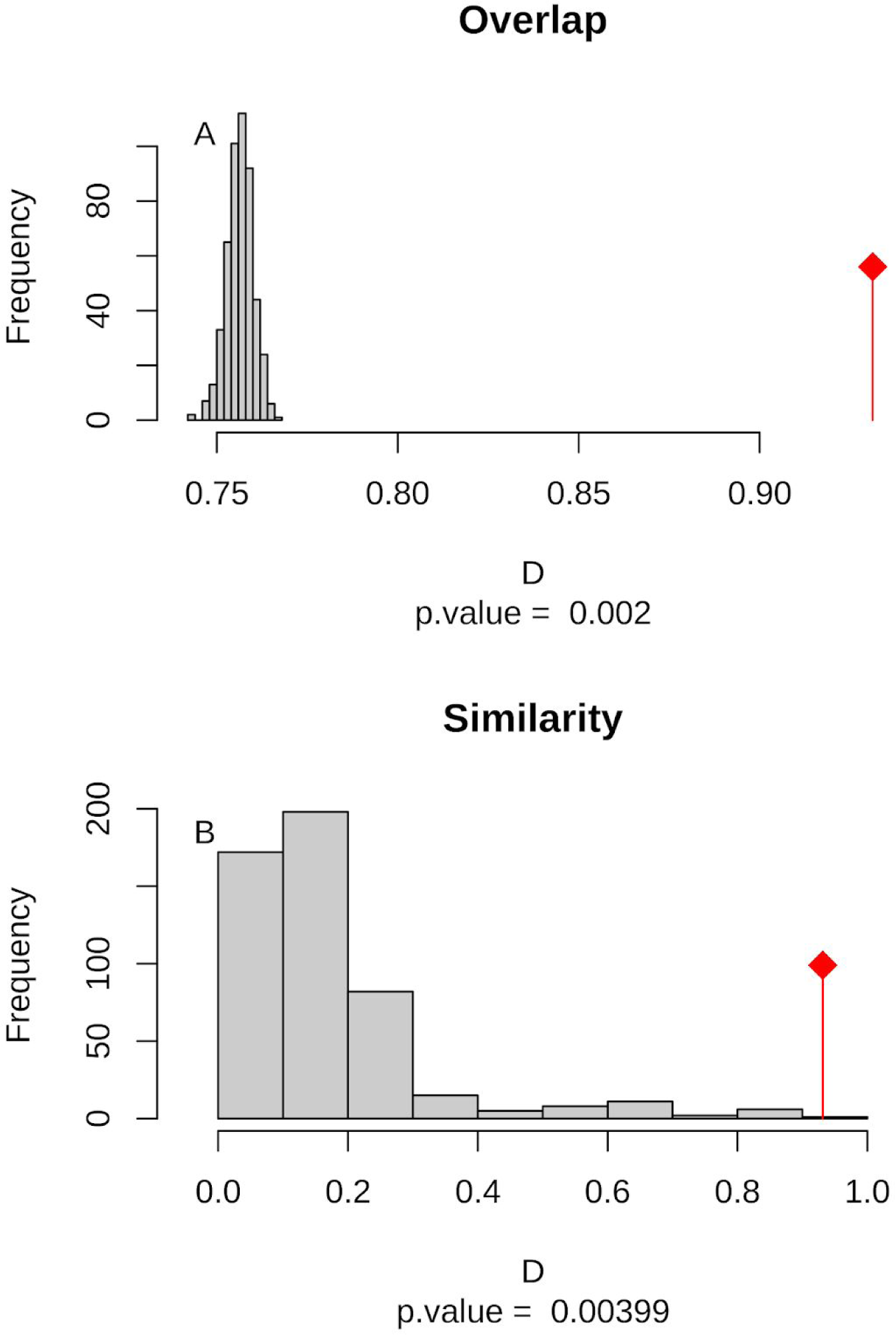
(A) Niche overlap and (B) similarity tests for Maxent species distribution models built with population scaled SARS-CoV-2 coronavirus data compared to one built with human population density as occurrence data; actual model overlap indicated by a red marker in both plots. Significant *p*-values correspond to greater niche overlap or similarity than expected by random models.

## Discussion

### Main findings

Our results suggest that population scaled SARS-CoV-2 coronavirus cases are highly linked with the human populations in the US, and that any influence of climate is currently hard to disentangle for SARS-CoV-2 cases in the US (**Figures 1-4**). Furthermore, this link is strengthening over time (**Figure 1**). This indicates that caution should be used when dealing with climate modeling of SARS-CoV-2, at least on a national scale. Our results, while broadly similar to what was found in the US for global scale models (Araújo & Naimi, 2020; Bariotakis et al., 2020), indicate that the pattern we observed is simply reflecting human population density. Please note that an updated online version (https://navaak.shinyapps.io/CVRisk/; accessed April 4, 2020) of the figure in one of these studies (Bariotakis et al., 2020) predicts a rather different model from their original (e.g., low suitability for much of the US). Based on this, at least some human-focused data (i.e., host data) should be compared or incorporated in any modeling exercises for SARS-CoV-2. Accordingly, the current global results from other studies (Araújo & Naimi, 2020; Bariotakis et al., 2020) should not be taken at face value without a critical comparison to human distributions.

It is also noteworthy that our models using the raw viral case data (**Figure S2**), as opposed to population scaled data (**Figure 3**), performed poorly: the raw SDM did not highlight most of the currently known range of SARS-CoV-2 in the US. Other SDM studies have only used raw values as inputs (Araújo & Naimi, 2020; Bariotakis et al., 2020), which our model, again, indicates should not be used. Similarly, our analysis of raw SARS-CoV-2 case data shows that over time the probability density curves focus on a narrower range of climate as more data comes from the areas most affected by the outbreak (**Figure S1**). In contrast, adjusting for population size with the population scaling results in SARS-CoV-2 climate curves more closely approaching that of the human population curves over time (**Figure 1**). Furthermore, county-level human population numbers only appear to be a strong correlate to raw cases, rather than population scaled cases (**Figure 2**). Together these observations suggest that the SARS-CoV-2 is highly successful in a few areas at this point (e.g., New York City), but when scaled for the available human population in each county we can infer a much broader geographic and climatic range available.

Again, our data have a few indications that climate may have some relevance in SARS-CoV-2 distributions. Viral probability densities are slightly higher in cooler locales (**Figure 1, S1**). The population scaled data are particularly compelling, even if the climate difference is smaller, as they are less driven by the current SARS-CoV-2 hotspots. Furthermore, our population scaled viral SDMs, while statistically identical to our human SDMs, have lower suitability for the virus in Florida and the southwestern border (**Figure 3**). In reality, Florida currently has a reasonably high number of viral cases.

### Avoiding a presumably faulty prediction

Given that our SDMs for humans and the virus were statistically indistinguishable, we do not believe that a future projection of our SARS-CoV-2 SDM in the US using only climate data would be trustworthy at this point in time, and therefore do not present one. The model likely would suggest the SARS-CoV-2 prevalence will shift northward during the summer, which would be congruent with a recent global modeling study (Araújo & Naimi, 2020). However, given that humans do not move northward in large numbers in the US during the summer, our predictions would be based on faulty presumptions about host resources (i.e., that there are sufficient humans in the north to harbor the bulk of cases). Still, it is possible that a future model projection would turn out to be correct, and there could be at least some residual predictive power that is not fully encompassed by human population patterns. It is our hope that as data improve we, and others, may be able to increase our confidence in, or better refute, SDM-based future projections and other climate-focused SARS-CoV-2 studies. Adding human population density as an input layer in an SDM might be a straightforward way to avoid these issues and produce a more trustworthy prediction.

### Modeling counterarguments

It has now been argued that SDM modeling of SARS-CoV-2 can help to predict where the virus may generally be found now and in the future (Araújo & Naimi, 2020; Bariotakis et al., 2020), as well as why it might be a fool’s errand to conduct these analyses in the first place (Chipperfield et al., 2020). Here are some of the main arguments that have been put forward against SARS-CoV-2 SDMs, followed by our opinion on the subject. 1) Issue: The virus is spreading and the population is not in equilibrium with climate or any other putative niche dimension. Our thoughts: It is not in equilibrium and that is imperfect; however, SDMs have been useful for a number of non-equilibrium systems, like species invasions during their spreading phase (Václavík & Meentemeyer, 2009), and waiting for equilibrium will mean predictions are no longer as useful to conduct (i.e., would only be helpful for a next outbreak of this virus). Furthermore, the rebuttal’s reanalysis of global virus cases (their Figure 2), in our opinion, seemed to indicate there may already be some stability in global model projections (Chipperfield et al., 2020); however, the rebuttal authors interpreted the relative model differences to claim instability and, therefore, issues with reliability. 2) Issue: The virus may be spreading heavily or underreported in the Global South. Our thoughts: Surely this is partially true, but this may not completely explain observable climatic correlations. Even if we are working with incomplete data, it is hard to know if adding those data will broadly change conclusions based on preliminary models. Still, to mitigate this problem we focused on the more consistent US-level data. 3) Issue: Papers must strive for best practices. Our thoughts: Yes, we agree that best practices are indeed worth pursuing, even if not everyone agrees on what best practices may be. We have done our best to achieve the somewhat aspirational standards that have been put forth (Araújo et al., 2019). We explicitly document this, and summarize shortcomings that are inherent with a newly spreading system (**Table 1**). 4) Issue: Caution should be taken in claims and dissemination of research that could impact public health policy. Our thoughts: Yes, caution is advisable in terms of studies on dire subjects like SARS-CoV-2; however, cross pollination between disciplines has been important for many breakthroughs and advances in science.

### Caveats and unknowns

There are individual people who regularly go undetected for SARS-CoV-2 (e.g., those who lack symptoms or have mild symptoms) — currently estimated at up to 25% (Mandavilli, 2020). This is of course an issue for all studies of SARS-CoV-2. Obviously it is not ideal for modeling and forecasting; however, it is inherent in any study. In fact, we have far more data available for SARS-CoV-2 than we will ever have for the vast majority of viruses and biodiversity generally.

Our results are based on data for a single, large country. While taking global data points would be ideal, we avoided these because they are known to be more problematic and inconsistent (Chipperfield et al., 2020). While it may somewhat hobble our ability to forecast viral distributions at other time points, as there are surely non-analogous weather systems to come (i.e., no place in the US is currently as hot as Death Valley, California in full summer heat), we feel it was a worthy trade-off. Policy, social factors, and a variety of other variables were not included in this study, despite their obvious, known, or potential importance (Leung et al., 2020; Maier & Brockmann, 2020; Miller et al., 2020); other variables such as wearing masks and prior Bacillus Calmette-Guérin childhood vaccinations may be important for global patterns (Leung et al., 2020; Maier & Brockmann, 2020; Miller et al., 2020). Unfortunately, data on these variables are often lacking in public databases. Our goals were to focus on a rather macro scale, which may be harder to do with policy data that can vary substantially from county to county, state to state, and, for a global study, country to country. While policies in the US may not have fully been implemented for long enough to tell, these changes (e.g., self isolation) will surely impact models in upcoming days.

### Data accessibility and mobilization

We believe the New York Times (The New York Times, 2020) providing robust county-level data in an accessible repository with an open license for the US sets an excellent standard that should be repeated by governments, academics, and other media organizations for other parts of the world so that this type of study may be better repeated in any country or globally. In a similar vein, we have made our analytical and modeling pipeline (along with figures) available (https://github.com/rsh249/cv19_enm/releases/tag/v0.0.2). We believe it is imperative for all pipelines and scripts to be made available for any SARS-CoV-2 research to ensure that models can be improved upon and any errors can be more quickly uncovered and resolved.

### Future directions

There are many avenues to pursue regarding SARS-CoV-2 modeling and predictions. We are excited to see researchers from a variety of fields extending their toolkits towards understanding this virus. We hope that ecological studies like this and others can play a role without overcomplicating the research efforts put forth by epidemiologists. Still, studies should familiarize themselves with current critiques of SDMs for SARS-CoV-2 modeling and be cautious of their inputs and conclusions.

With improving data, we feel that future studies should better be able to examine the system globally while considering human populations and public policy efforts at curbing the virus. We also believe that it will at some point in the US and elsewhere be worth examining death rates in different areas, as it would be helpful to know if climate or other abiotic variables might impact this heartbreaking statistic. Coupling regularly updated data with automated online resources would also be particularly helpful in learning how this virus may spread.

### Conclusions

At present, SDMs from SARS-CoV-2 population scaled cases do not appear to be distinguishable from human population density. It may be too early to tell if climate will play a significant role for this coronavirus in the US with warmer months approaching. Future studies looking at climate’s impact on this virus should, wherever possible, take into account human population density in any analyses.

## Data Availability

Data and code for this study are archived at https://github.com/rsh249/cv19_enm/releases/tag/v0.0.2

## Acknowledgements

We thank those who are treating the sick, providing essential services, preventing viral spread, sheltering alone, and helping others during this time of need; we thank the New York Times for making their data publically available; we thank the other researchers who have been providing a lively debate on best practices and how to make predictions at a point in time when our methods may matter most; and we thank Peter Galante and Rob DeSalle for reviewing a draft of this manuscript.

## Supplementary Information

**Figure S1.**
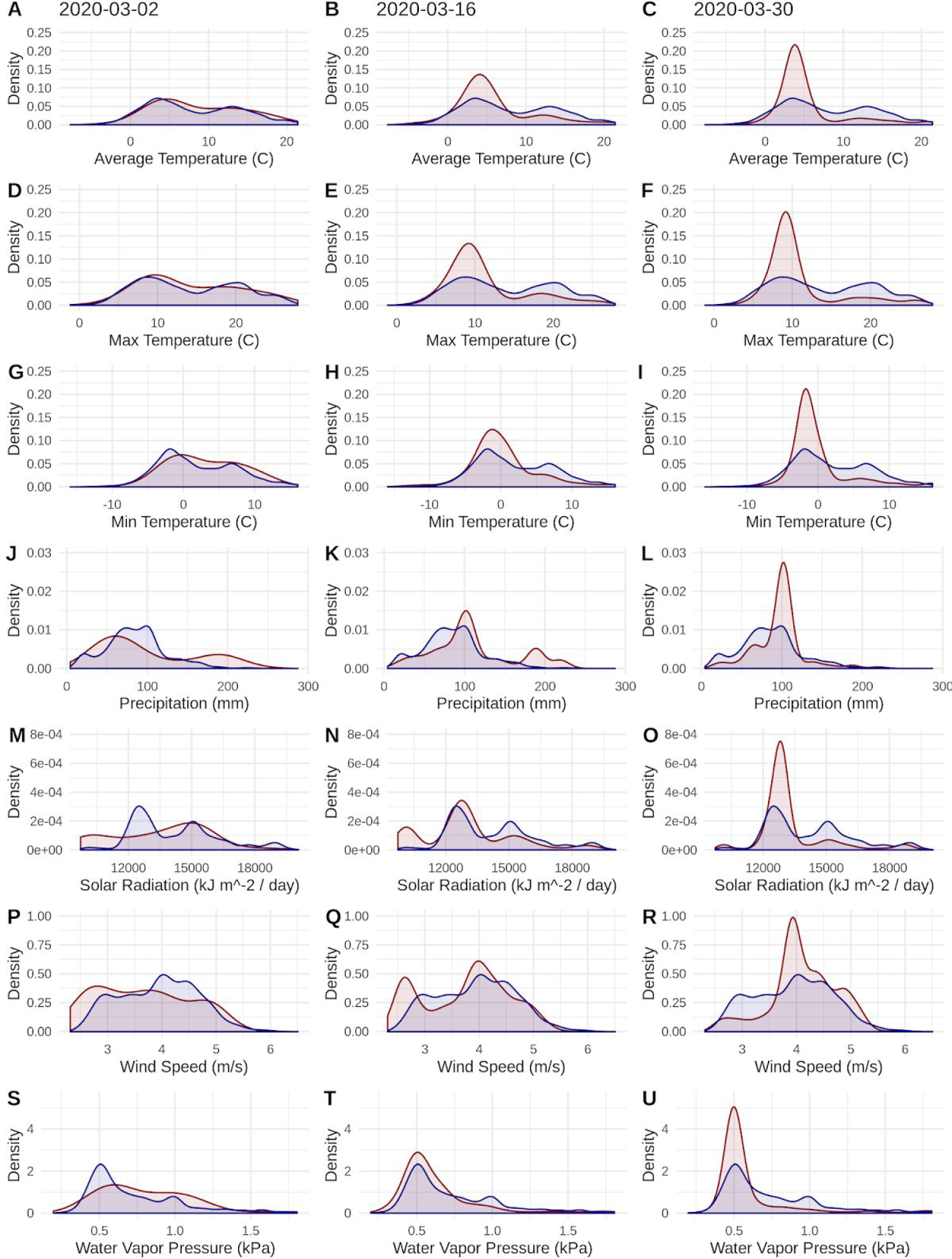
Equivalent of Figure 1, but using raw virus data. Probability densities of SARS-CoV-2 coronavirus cases (using raw data; curves in red) compared to the probability densities of human populations (curves in blue) in each US county for each of seven climate variables. Probability density curves are standardized to an area of one.

**Figure S2.**
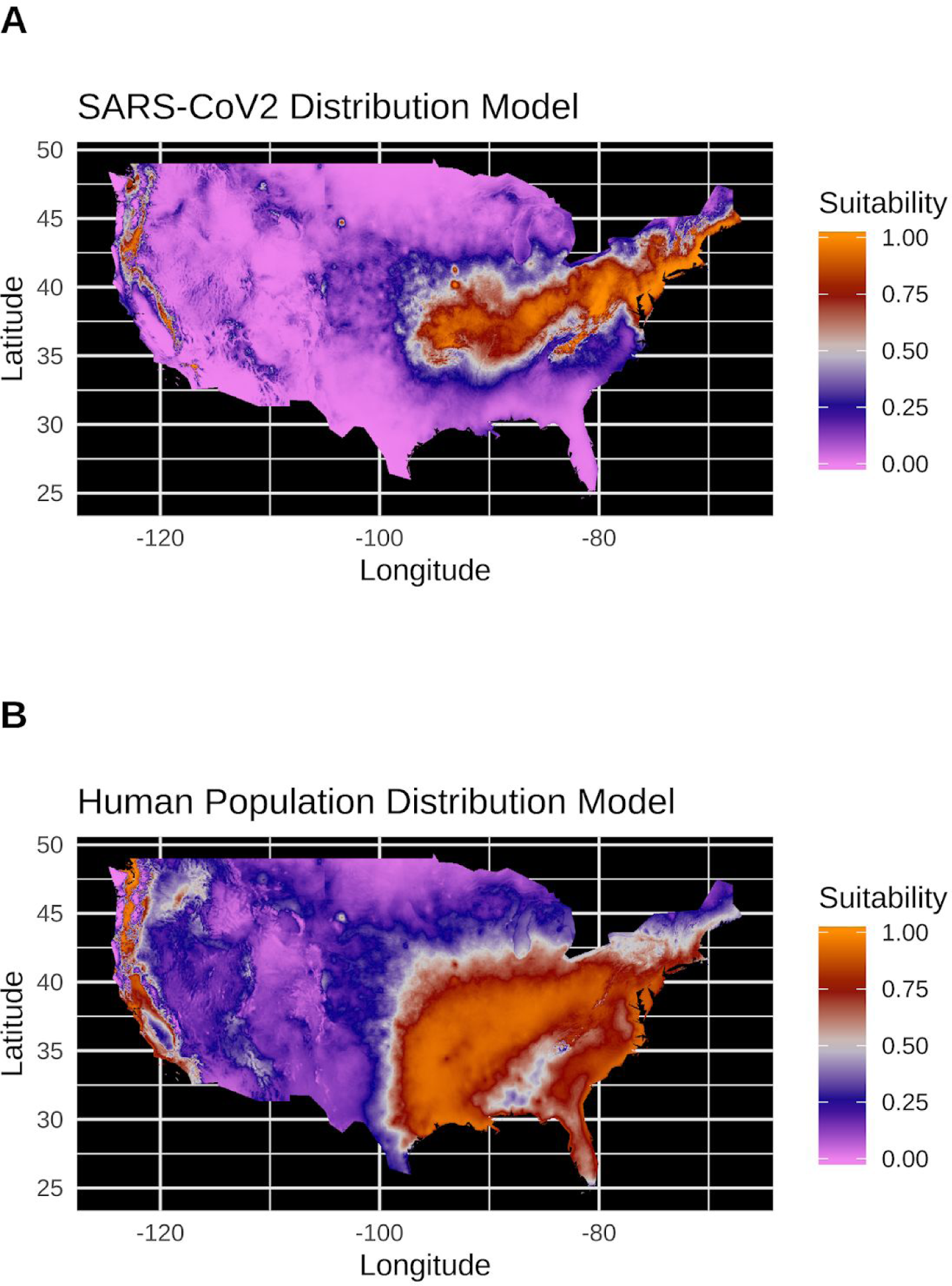
Equivalent of Figure 3, but using raw virus data. (A) Species distribution model of the SARS-CoV-2 coronavirus (using raw data) for March 30, 2020. (B) Human population distribution model for the US from 2010.

**Figure S3.**
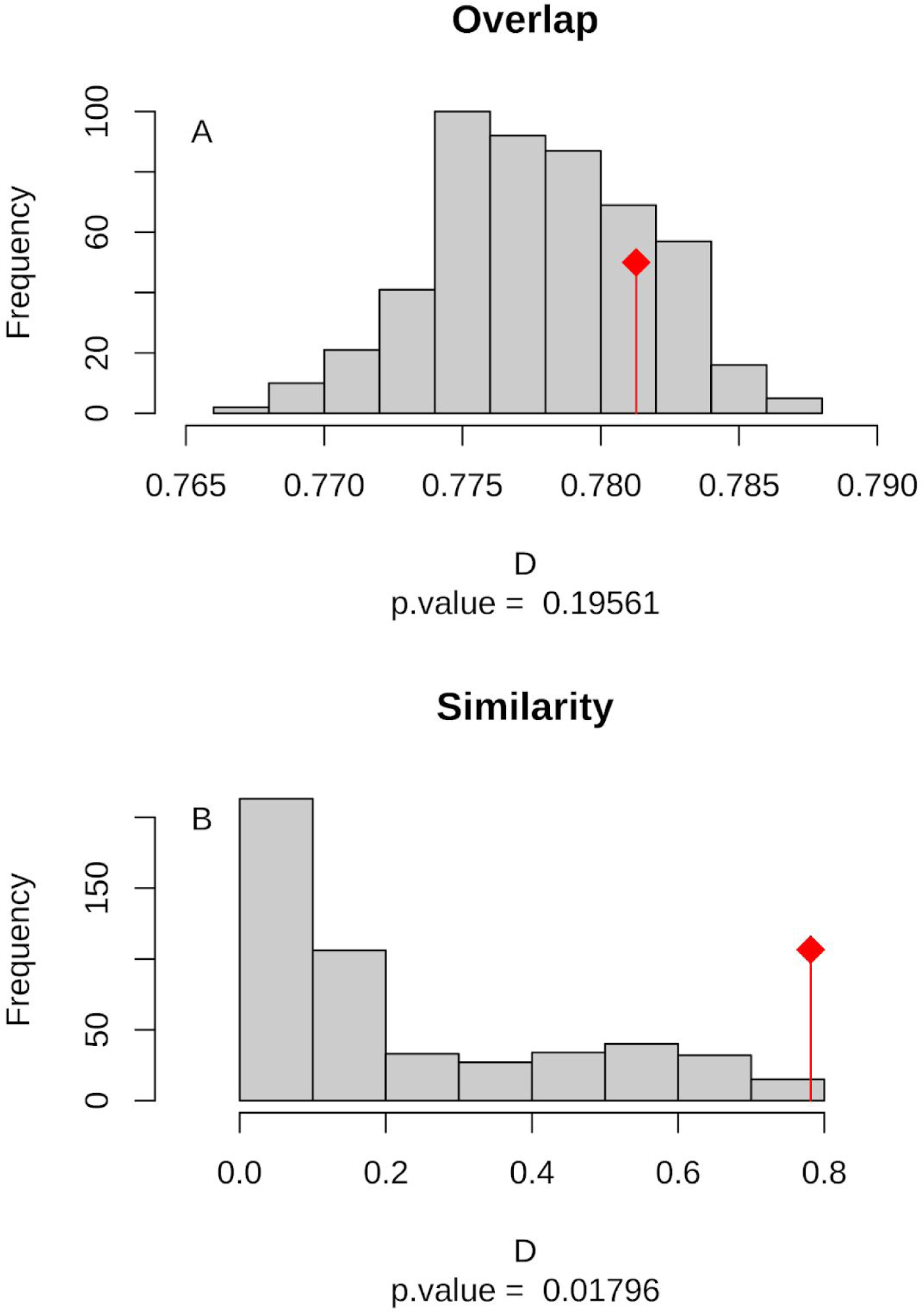
Equivalent of Figure 4, but using raw virus data. (A) Niche Overlap and (B) similarity tests for Maxent species distribution models built with SARS-CoV-2 coronavirus case data compared to one built with human population density as occurrence data; actual model overlap indicated by a red marker in both plots. Significant *p*-values correspond to greater niche overlap or similarity than expected by random models.

